# Chronotropic Incompetence among People with HIV Improves with Exercise Training in the Exercise for Healthy Aging Study

**DOI:** 10.1101/2023.11.10.23298367

**Authors:** Matthew S. Durstenfeld, Melissa P. Wilson, Catherine M. Jankowski, Grace L. Ditzenberger, Chris T. Longenecker, Kristine M. Erlandson

## Abstract

**Background:** People with HIV (PWH) have lower exercise capacity compared to HIV uninfected peers, which may be explained by chronotropic incompetence (CI), the inability to increase heart rate during exercise.

**Methods:** The Exercise for Healthy Aging Study included adults ages 50-75 with and without HIV. Participants completed 12 weeks of moderate intensity exercise, before randomization to moderate or high intensity for 12 additional weeks. We compared adjusted heart rate reserve (AHRR; CI <80%) on cardiopulmonary exercise testing by HIV serostatus, and change from baseline to 12 and 24 weeks using mixed effects models.

**Results:** Among 32 PWH and 37 controls (median age 56, 7% female, mean BMI 28 kg/m_2_), 28% of PWH compared to 11% of controls had CI at baseline (p=0.067). AHRR was lower among PWH (91 vs 102%; difference 11%, 95% CI 2.5-19.7; p=0.01). At week 12, AHRR normalized among PWH (+8%, 95% CI 4-11; p<0.001) and was sustained at week 24 (+5, 95%CI 1-9; p=0.008) compared to no change among controls (95%CI –4 to 4; p=0.95; p_interaction_=0.004). After 24 weeks of exercise, only 15% PWH and 10% of controls had CI (p=0.70).

**Conclusions:** Chronotropic incompetence contributes to reduced exercise capacity among PWH and improves with exercise training.

## Introduction

People with HIV (PWH) have excess cardiovascular disease [1] and reduced cardiorespiratory fitness (peak exercise capacity) compared to the general population without HIV [2–7]. A major contributor to reduced cardiorespiratory fitness among PWH may be chronotropic incompetence, which occurs in approximately one third of PWH [8, 9]. Chronotropic incompetence is the inability to increase heart rate adequately during exercise to meet increased metabolic demands, which results in a lower peak exercise capacity. The gold-standard test for chronotropic incompetence is cardiopulmonary exercise testing (CPET) which includes measurement of gas exchange to quantify exercise capacity in terms of oxygen consumption (peak VO_2_). Among people without HIV, chronotropic incompetence even among asymptomatic individuals without cardiovascular disease is associated with increased risk of development of incident cardiovascular disease and mortality [10–14]. The two prior studies of chronotropic incompetence among PWH were both cross-sectional and did not include assessments of body composition or baseline physical activity which could be unmeasured confounders in the observed association between HIV and chronotropy [8, 9].

Little is known about treatment of chronotropic incompetence outside of populations of people with heart failure. In that patient population, implanting pacemakers to increase maximal exertional heart rate does not improve symptoms or exercise capacity [15]. In contrast, supervised exercise training improves chronotropy and exercise capacity among people with heart failure [16]. Observational data among people with pacemakers suggests that improvements in chronotropy are associated with improved quality of life [17]. However, whether a supervised exercise training program modifies chronotropic incompetence among people without heart failure, especially among PWH, has not been studied to our knowledge.

Therefore, we designed this secondary analysis of 75 the Exercise for Healthy Aging Study, a clinical trial of a supervised exercise training intervention among people with and without HIV, to test the hypotheses that (1) HIV would be associated with chronotropic incompetence at baseline, and (2) chronotropy would improve with exercise training.

## Methods

### Study Design

This study is a secondary analysis of the Exercise For Healthy Aging Study, a clinical trial of an exercise intervention among sedentary adults age 50-75 with and without HIV from 2014-2017 (clinicaltrials.gov NCT02404792).[18] Participants exercised at moderate intensity for weeks 1-12 and were then randomized to continue moderate or advance to a higher intensity of exercise for weeks 13-24.

### Participants

The Exercise for Healthy Aging study enrolled people with HIV (PWH) and HIV uninfected controls age 50-75 years who were sedentary (<60 minutes of physical activity each week for 6 months by self-report) and without contraindications to participating in an exercise program; PWH were required to be on stable antiretroviral therapy with viral load < 200 copies/mL and CD4 count greater than 200 cells/μl. Full inclusion and exclusion criteria have been published [18].

### Study Procedures

*Cardiopulmonary Exercise Testing:* All participants underwent a symptom-limited graded CPET using a treadmill at baseline, 12 weeks, and 24 weeks as previously described [18]. The speed was adjusted to initially target 70% of the age-predicted maximum heart rate with the grade increased by 2% every 2 minutes until volitional exhaustion or test termination for safety. Participants who did not initially reach 85% of maximum predicted heart rate on the baseline study were required to undergo cardiology consultation prior to proceeding, and one person received revascularization and then was subsequently rescreened.

*Exercise Training Intervention:* Details of the intervention have been previously published [18]. Briefly, participants attended in-person, supervised exercise training program 3 times weekly for 24 weeks. The program started with 2 weeks of low-intensity exercise followed by 10 weeks of moderate-intensity exercise including both aerobic endurance exercise performed on a treadmill and strength training. At 12 weeks, participants underwent repeat CPET and were randomized to continue moderate intensity exercise or advance to high-intensity for an additional 12 weeks. Following completion of the 24-week program, participants underwent repeat CPET.

*Primary Exposure:* The primary exposure of interest was HIV infection.

*Primary Outcomes:* The primary outcomes of interest were the adjusted heart rate reserve (AHRR), a measure of exertional chronotropy defined by AHRR=(HR_peak_-HR_rest_)/(220-Age-HR_rest_). Chronotropic incompetence as a binary variable was defined by an adjusted heart rate reserve <80%. Participants on chronotropic medications (beta-blockers, calcium channel blockers) were considered to have chronotropic incompetence if AHRR<62% [19, 20]. The primary longitudinal outcomes were change in AHRR and chronotropic incompetence at 12 weeks and 24 weeks.

*Secondary Outcomes:* The main secondary outcome was exercise capacity as measured by the peak oxygen consumption (Peak VO_2_) during CPET.

*Covariates:* We included age, sex, body mass index, body composition variables assessed using dual energy x-ray absorptiometry (DEXA) as previously reported [21], hypertension, diabetes, and HIV specific factors including nadir and current CD4^+^ cell count, years with diagnosis with HIV, years of treatment with antiretroviral therapy (ART), and current and prior ART medications. Baseline physical activity was assessed using the International Physical Activity Questionnaire short form [22]. Biomarkers including high sensitivity C-reactive protein (hsCRP), interleukin 6 (IL-6), interleukin 10 (IL-10), tumor necrosis factor (TNF), soluble TNF receptor 1 (sTNFR-1), soluble TNF receptor 2 (sTNFR-2), soluble CD14 (sCD14), and insulin-like growth factor binding protein 3 (IGFB-3) were measured on baseline plasma samples using commercially available enzyme-linked immunosorbent assays as previously reported [23].

*Statistical Analysis:* Study data were collected and managed using REDCap. A deidentified dataset was generated and provided for this secondary analysis. AHRR at baseline was compared using t-tests for unadjusted analyses and linear regression with HIV as the primary predictor of interest and including age, sex, and body mass index as pre-specified covariates with other covariates of interest includes in secondary analyses. Chi squared tests were used to compare the proportion with and without chronotropic incompetence by HIV status. Factors associated with chronotropic incompetence including HIV specific factors were reported by univariate analysis (Fisher’s exact/chi-squared test for binary, t-test or Pearson correlation coefficients for AHRR). We subsequently included all of the cardiovascular risk factors in a linear regression model. For the longitudinal analyses, we used linear mixed effects models with random slopes and intercepts by participant, an independent covariance structure, and a clustered sandwich estimator. We considered p<0.05 to be significant for the primary outcomes and did not adjust for multiple comparisons. Given the small sample size, we only assessed for interactions with the physical activity and body composition variables and HIV status. Analyses were performed using STATA 17.0.

*Approval:* The original study was approved by the Colorado Multiple Institution Review Board and written informed consent was obtained.

## Results

At baseline, 32 PWH and 37 controls completed CPET; 28 PWH and 31 controls completed 12 weeks of moderate-intensity exercise training and 27 PWH and 29 controls completed the full 24-week trial [18]. Participants had a median age of 56 years and were predominantly male (Table 1). Mean BMI was 28 kg/m^2^. Among PWH, median duration with diagnosed HIV was 21 years with a median treatment duration with antiretroviral therapy of 17 years.

**Table 1.** Participant Characteristics by HIV Status.

|  | HIV (n=32) | No HIV (n=37) | p value |
| --- | --- | --- | --- |
| Age | 55.5 (52, 60.5) | 58 (53, 64) | 0.34 |
| Female | 4 (13%) | 2 (5%) | 0.41 |
| Male | 28 (88%) | 35 (95%) | 0.54 |
| Race |  |  | 0.09 |
| Black | 9 (28%) | 3 (8%) |  |
| White | 20 (63%) | 31 (84%) |  |
| More than 1 race or other | 3 (9%) | 3 (8%) |  |
| Hispanic/Latino | 4 (13%) | 4 (11%) | 1.00 |
| Medical History |  |  |  |
| Hypertension | 16 (50%) | 25 (68%) | 0.14 |
| Diabetes | 2 (6%) | 4 (11%) | 0.50 |
| Dyslipidemia | 14 (44%) | 22 (59%) | 0.19 |
| Depression | 17 (53%) | 9 (24%) | 0.01 |
| Coronary Artery Disease* | 1 (3%) | 2 (5%) | 1.00 |
| Veterans Aging Cohort Study Index, % $\geq 20$ points | 18 (56%) | 9 (24%) | 0.01 |
| Tobacco Use |  |  | 0.07 |
| - Never | 11 (34%) | 23 (62%) |  |
| - Smoked <10 years and quit | 4 (13%) | 5 (14%) |  |
| - Smoked >10 years and quit | 12 (38%) | 5 (14%) |  |
| - Current Smoker | 5 (16%) | 4 (11%) |  |
| Marijuana Use (in last year) | 19 (59%) | 5 (14%) | <0.001 |
| Alcohol Use |  |  | 0.21 |
| None | 10 (31%) | 6 (16%) |  |
| 2x/week or less | 19 (59%) | 23 (62%) |  |
| Daily | 3 (9%) | 8 (22%) |  |
| Body Composition |  |  |  |
| Body Mass Index, kg/m<sup>2</sup> | 25.0 (23.8, 28.4) | 28.3 (26, 32.3) | 0.01 |
| Total Lean Mass, kg | 58.7 (51.6, 64.2) | 59.7 (55.8, 69.1) | 0.06 |
| Total Fat Mass, kg | 20.0 (15.3, 24.0) | 26.3 (22.5, 33.5) | 0.005 |
| Visceral Adipose Tissue Area, cm<sup>2</sup> | 171 (133, 227) | 219 (173, 261) | 0.09 |
| Appendicular Lean Mass, kg/m<sup>2</sup> | 25.9 (22.6, 29.5) | 26.3 (25.1, 31.4) | 0.07 |
| Percent Fat | 29±7 | 33±7 | 0.02 |
| Physical Activity (IPAQ) |  |  |  |
| Days/Week >10 minutes of Vigorous Activity | 0 (0, 2) | 0 (0, 1) | 0.35 |
| Days/Week >10 minutes Moderate Activity | 1 (0, 2) | 1 (0, 2) | 0.19 |
| Days/Week >10 minutes Walking | 3 (0, 7) | 5 (1, 7) | 0.69 |
| MET minutes per week | 863 (56, 2097) | 1215 (264, 2856) | 0.77 |
| Awake Hours Sitting per Day | 6 (4, 9) | 6 (4,9) | 0.61 |
| Short Physical Performance Battery |  |  | 0.14 |
| Score |  |  |  |
| Score≥12 (no impairment) | 21 (66%) | 31 (84%) | |
| Score<12 | 11 (34%) | 6 (16%) |  |
| Current Statin Use | 13 (41%) | 15 (41%) | 1.00 |
| Integrase inhibitor-containing regimen | 21 (66) |  |  |
| Protease inhibitor-containing regimen | 13 (41) |  |  |
| NNRTI-containing regimen | 9 (28) |  |  |
| Any prior zidovudine treatment | 12 (38) |  |  |
| Any prior stavudine treatment | 14 (44) |  |  |
| Current CD4<sup>+</sup> cell count | 591 (449, 836) |  |  |
| Years since HIV diagnosis | 21 (16, 28) |  |  |
| Estimated years of continuous ART | 17 (6, 19) |  |  |
| Baseline Inflammatory Markers |  |  |  |
| C-Reactive Protein | 2.0 (1.0, 3.8) | 1.1 (0.7, 2.4) | 0.05* |
| IL-6 | 2.1 (1.4, 2.6) | 1.8 (1.4, 2.6) | 0.58* |
| IL-10 | 4.4 (3.2, 6.0) | 4.1 (3.0, 7.9) | 0.95* |
| TNF-α | 1.3 (1.1, 1.7) | 1.0 (0.8, 1.3) | 0.01* |
| sTNFR1 | 1147 (937, 1488) | 1062 (904, 1196) | 0.18* |
| sTNFR2 | 2769 (2078, 4111) | 2289 (1863, 2865) | 0.05* |
| sCD14 | 1779 (1595, 1959) | 1596 (1309, 1691) | 0.004* |
| IGFBP-3 | 1704 (1102, 2231) | 1931 (1552, 2297) | 0.13* |

\*Wilcoxon rank sum
Table Legend: Coronary artery disease was defined as prior myocardial infarction, bypass surgery, angioplasty, or stent. IL=interleukin. TNFR=tumor necrosis factor receptor. CD=Cluster of Differentiation. IGFBP=insulin like growth factor binding protein.

### Chronotropy Prior to Exercise Training

Baseline CPET results are presented in Table 2. At baseline, mean AHRR was 91% (95%CI 85-96) among PWH compared to 101% among controls (95%CI 95-108; unadjusted difference 10.5%, 95%CI 2.5-19.7; p=0.02; Figure 1). With adjustment for age, sex, and body mass index, AHRR was 13% lower among PWH (95% CI 3.8-22.4; p=0.007). In a sensitivity analysis excluding those with coronary artery disease or on chronotropic medications at baseline (n=4), the magnitude of the association remained unchanged (13% lower among PWH; 95%CI 5-21; p=0.002). Among PWH, 28% had chronotropic incompetence compared to 11% of controls (OR 3.2; 95%CI 0.89-11.8; p=0.08).

**Figure 1.**
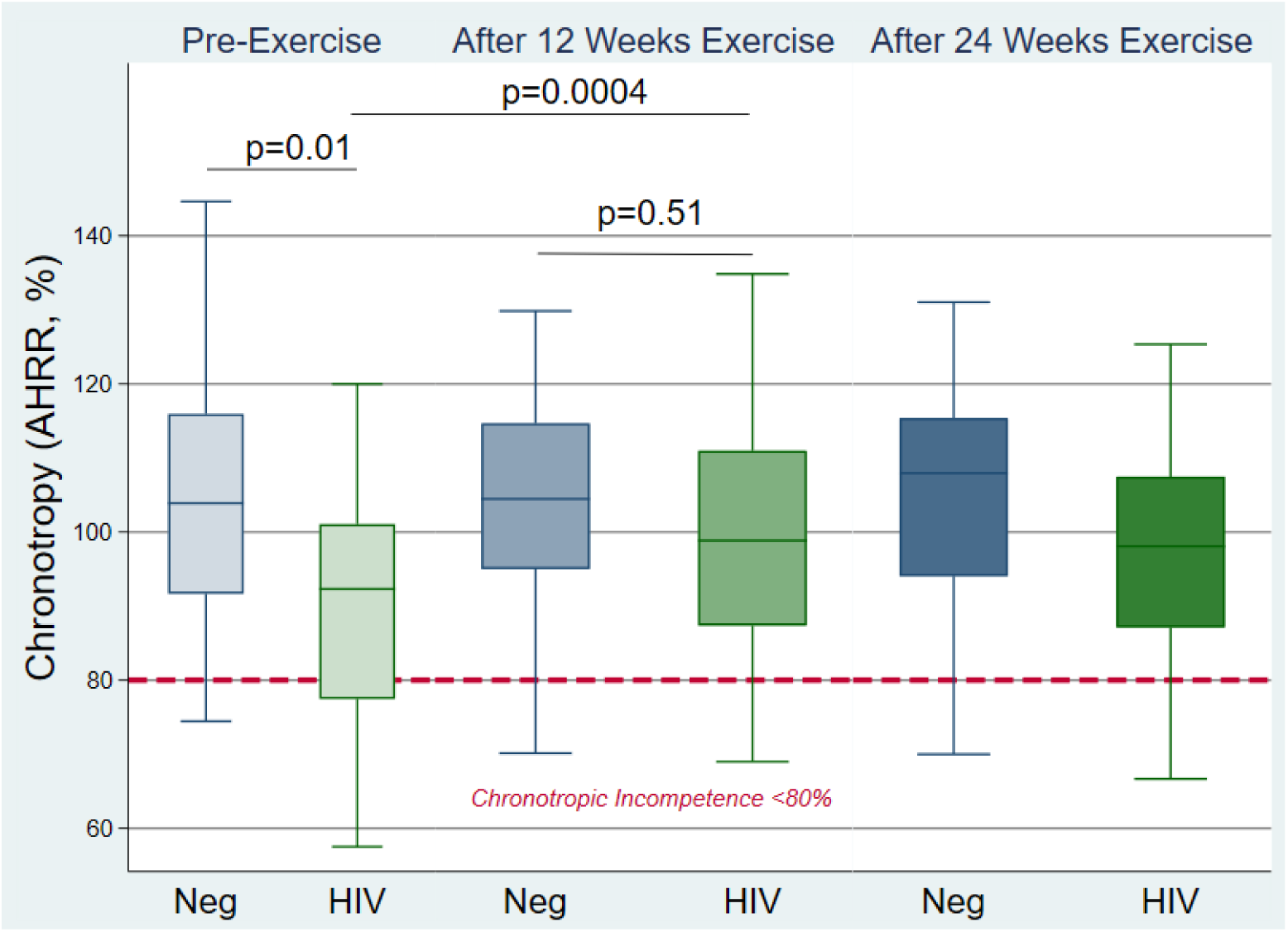
Chronotropy by HIV Status and Time. Figure Legend: Boxplots (median, interquartile range) for adjusted heart rate reserve among those with and without HIV at baseline, after 12 weeks, and after 24 weeks of exercise. Dashed line represented the definition for chronotropic incompetence among people not on chronotropic medications (<80%).

**Table 2.** Baseline Cardiopulmonary Exercise Testing Results by HIV.

|  | PWH (n=32) | Controls (n=37) | P value |
| --- | --- | --- | --- |
| Duration of Exercise | 10.1±3.2 minutes | 9.9±2.6 minutes | 0.78 |
| Respiratory Exchange Ratio<sup>a, b</sup> | 1.09±0.07 | 1.09±0.6 | 0.71 |
| Peak VO<sub>2</sub> (ml/kg/min)<sup>b</sup> | 25.9±5.7 | 27.2±6.1 | 0.35 |
| Resting (standing) heart rate | 75±10 | 79±10 | 0.09 |
| Maximum heart rate | 155±13 | 163±17 | 0.08 |
| Age-Predicted Maximum heart rate, % (220-age) | 95±8 | 100±10 | 0.02 |
| Adjusted heart rate reserve, % | 88±21 | 101±20 | <0.01 |
| Chronotropic Incompetence, n (%) | 9 (28%) | 4 (11%) | 0.07 |

Footnotes: a: 3 baseline studies among PWH were submaximal. b. One baseline study was missing gas exchange information.

At baseline, AHRR was correlated with peak VO_2_ (ρ=0.45, p=0.0001; Figure 2). Chronotropic incompetence at baseline was associated with 5.3 ml/kg/min lower peak VO_2_ (95% CI –8.1 to –2.5; p<0.001) adjusted for age, sex, and body mass index. Each 10% decrease in AHRR was associated with a 1.4 ml/kg/min reduction in peak VO_2_ (95% CI –2.0 to –0.8; p<0.001).

**Figure 2.**
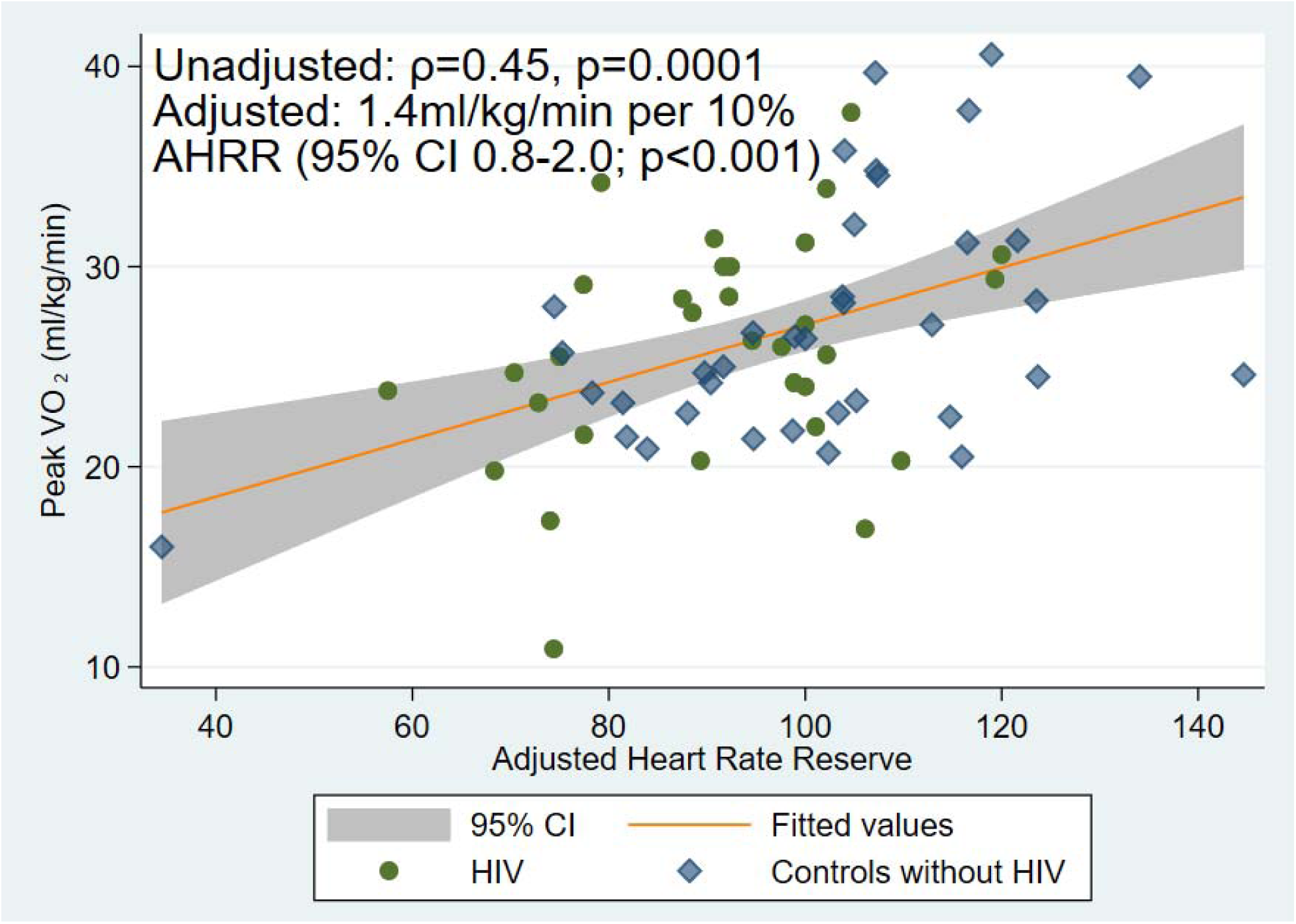
Association between Chronotropy and Cardiorespiratory Fitness. Figure 2 Legend: Scatter plots and linear fit of the association between adjusted heart rate reserve (x-axis) and peak VO_2_ in ml/kg/min at baseline. Adjusted for age, sex, and body mass index, each 10% increase in AHRR was associated with a 1.4 ml/kg/min higher peak VO_2_.

### Factors Associated with Baseline Chronotropy

In exploratory analyses, we found that cardiovascular risk factors at baseline were associated with chronotropic incompetence. In univariate analysis, hypertension, coronary artery disease, chronotropic medications, current smoking, but not diabetes or dyslipidemia, were correlated with lower AHRR. As shown in Table 3, in a model including age, sex, body mass index, hypertension, dyslipidemia, diabetes, current smoking, coronary artery disease, and use of chronotropic medications, coronary artery disease was associated with 29% lower AHRR (95%CI 10-47; p=0.004) and chronotropic medications with 25% lower AHRR (95% CI 2-48; p=0.03). HIV remained significantly associated with chronotropy even accounting for coronary artery disease and chronotropic medications (9% lower AHRR; 95% CI 1-17; p=0.03). Other factors are listed in Table 3.

**Table 3.** Factors Associated with Chronotropic Incompetence.

|  | Adjusted Difference in AHRR | 95% Confidence Interval | P value |
| --- | --- | --- | --- |
| HIV<sup>a</sup> | -13% | -22, -4 | <0.01 |
| Hypertension<sup>b</sup> | -10% | -18, -1 | 0.02 |
| Dyslipidemia<sup>b</sup> | +7% | -2, 15 | 0.06 |
| Diabetes<sup>b</sup> | +4% | -10, 17 | 0.93 |
| Smoking<sup>b</sup> | -16% | -27, -6 | 0.03 |
| Coronary Artery Disease<sup>b</sup> | -29% | -47, -10 | <0.01 |
| Chronotropic Medication Prescribed<sup>b</sup> | -25% | -48, -2 | 0.03 |
| Body Mass Index<sup>c</sup> | -17% per standard deviation increase | -11, -2 | 0.03 |
| Visceral Adipose Tissue Area<sup>c</sup> | -6.6% per standard deviation increase | -11, -2 | <0.01 |
| Visceral Adipose Tissue Area Independent of BMI<sup>d</sup> | -3.9% per standard deviation increase | -11.6, 3.8 | 0.14 |
| Baseline Physical Activity (MET-Hours per week)<sup>c</sup> | 0.04 per MET-hour/week | -0.09, 0.02 | 0.49 |
| IL-6<sup>c</sup> | -6% per doubling | -10, -1 | 0.02 |

Footnotes: a. Adjusted for Age, Sex, Body Mass Index. b. Adjusted for Age, Sex, Body Mass Index, hypertension, dyslipidemia, diabetes, smoking, coronary artery disease, and chronotropic medications prescribed. C. Adjusted for Age, Sex, and HIV. D. Adjusted for Age, Sex, Body Mass Index, and HIV.

Some body composition parameters were modestly correlated with chronotropy. In univariate analysis, body mass index (ρ=-0.27 p=0.03) and visceral adipose tissue (VAT) area (ρ=-0.26, p=0.03) were negatively correlated with AHRR, with adjusted estimates reported in Table 3. Measures of lean body mass were not correlated with AHRR. There were negligible changes in estimates for the effect of HIV in models including DEXA-derived body composition variables individually or all together compared to models with BMI only or for VAT area substituted for BMI. As BMI and VAT are co-linear (ρ=0.79), both were associated with AHRR only when singularly included in the model (Table 3), with no interaction with BMI or VAT by HIV status (p_interaction_=0.80 and 0.44, respectively).

In contrast, among this sedentary population, baseline self-reported physical activity (in MET-hours per week) was not correlated with AHHR (ρ=-0.16, p=0.21). In an adjusted model including age, sex, body mass index and HIV, baseline physical activity was not associated with AHRR (β=-0.04 per MET-hour per week; 95%CI –0.09-0.02; p=0.20) with no interaction by HIV status (p_interaction_=0.87).

In univariate analysis restricted to PWH, current CD4 count and self-reported nadir CD4 were negatively correlated with AHRR (ρ=-0.22, p=0.04 and ρ=-0.23, p=0.04, respectively). Longer ART duration was positively correlated with higher (better) AHRR (ρ=0.22, p=0.046). In models including age, sex, body mass index, and HIV specific variables either individually or all together, there were no statistically significant factors identified.

Among the inflammatory markers measured, only IL-6 was associated with chronotropy. Baseline IL-6 was associated with a 7% lower AHRR per doubling (95% CI –13 to –1; p=0.02) accounting for age, sex, and BMI, which was attenuated when additionally accounting for HIV (5%; 95% CI –11-0.3; p=0.06). Other inflammatory markers including hsCRP, TNF, IL-10, sTNFR-1, sTNFR-2, sCD14, or IGFB-3 were not associated with chronotropy.

### Chronotropic Response to Exercise Training varies by HIV Status

At week 12, AHRR normalized among PWH (+8%, 95% CI 4-11; p<0.001) compared to 0.1% increase among controls (95% CI –4 to 4; p=0.95; p_interaction_=0.004: Figure 1). Mean AHRR at 12 weeks was 98% (95% CI 92-104) among PWH compared to 101% (95% CI 94-109) among controls (p=0.51). The improvement in chronotropy among PWH was sustained at week 24 (+5%, 95%CI 1-9; p=0.008). At week 24, mean AHRR was 97% (95%CI 91-103) among PWH and 104% (95%CI 98-110) among controls (p=0.09). Randomized assignment to the high intensity exercise intervention during weeks 12-24 was not associated with a difference in AHRR from 12 to 24 weeks compared to moderate intensity (p=0.84).

After 12 weeks of moderate intensity exercise 15% of PWH and 13% of controls met the definition for chronotropic incompetence (p=1.00). Similarly, after 24 weeks of exercise, 15% of PWH and 10% of controls, respectively, met the definition for CI (p=0.70). There was no difference at 24 weeks by intensity arm (p=0.39).

Each 10% increase in AHRR from baseline was associated with a 1.2 ml/kg/min increase in peak VO_2_ at 12 weeks (95% CI 0.5-1.8; p=0.001). Accounting for change in AHRR, the greater benefit of exercise observed among PWH at 12 weeks (peak VO_2_ increased +1.5 ml/kg/min more than controls, 95%CI 0.1 to 2.9, p=0.04) was attenuated (0.4 ml/kg/min, 95%CI –0.9 to 1.8; p=0.53).

## Discussion

Consistent with the existing literature, we found that chronotropic incompetence is a mechanism of reduced exercise capacity with a high prevalence among PWH. We expand upon the existing literature by focusing on a population of older sedentary adults with additional measures of body composition and physical activity. Adjusting for these baseline characteristics, our findings suggest that chronotropy among older sedentary PWH is not largely driven by the amount of baseline physical activity or body composition—that is, chronotropic incompetence is not likely to be simply a result of deconditioning. Importantly, we also provide the first evidence that chronotropic incompetence is modifiable in this patient population and that exercise training may be an effective intervention.

Research on chronotropic incompetence has only included cross-sectional assessments of chronotropy, with the exception of clinical trials in the setting of heart failure. To our knowledge, this is the first study to demonstrate that chronotropic incompetence is modifiable among patients without heart failure. Consistent with the literature in heart failure, this study provides evidence that exercise training improves chronotropy. A randomized clinical trial including individuals with heart failure with a reduced ejection fraction found that exercise training improved chronotropy and exercise capacity in the group assigned a supervised exercise training program compared to the control group [16]. There are no published studies testing exercise training or any other interventions for chronotropic incompetence among people without heart failure. Exercise training may improve chronotropy by decreasing baseline adrenergic activation seen in heart failure [16]. This leads to improved β-receptor responsiveness such that there is a greater increase in heart rate for a similar change in plasma catecholamines released during exercise. Another hypothesis is that exercise training might reduce inflammation. However, in this study, changes in inflammation were variable and did not consistently decrease [23]. Another hypothesis is that exercise improves mitochondrial efficiency and vascular function, allowing individuals to reach a higher maximal heart rate even if there is no change in the underlying β-receptor response.

Why the prevalence of chronotropic incompetence is higher among PWH is unclear. One interesting hypothesis-generating finding is that longer duration of ART is associated with better chronotropy; in earlier eras of diagnosis, lower nadir CD4 counts resulted in earlier treatment with ART. A likely contributor to chronotropic incompetence among people with successfully treated HIV may be increased chronic inflammation, which is associated with chronotropic incompetence in this study and prior work. Besides inflammation, prior papers have not identified HIV-specific factors that are clearly linked to chronotropic incompetence [8, 24]. Among people without heart failure, decreased β-receptor responsiveness is a likely mechanism of chronotropic incompetence [25, 26]. Chronic adrenergic activation from chronic inflammation that is present among PWH, even those who are treated with ART and virally suppressed, may be a major factor driving both chronotropic incompetence and heart failure with preserved ejection fraction among PWH [27–29]. Although a number of inflammatory markers were increased among PWH in this study, only IL-6 was inversely associated with chronotropy among people with and without HIV, consistent with prior reports [24, 30]. Ultimately, chronic inflammation may lead to cardiac interstitial fibrosis which is increased among PWH [31] but not yet strongly linked to chronotropic incompetence.

Our finding that cardiometabolic risk factors and smoking are associated with chronotropic incompetence is consistent with prior work among people without HIV [30, 32–36]. Although PWH have a higher burden of cardiovascular risk factors, these do not fully explain differences in baseline chronotropic incompetence by HIV serostatus. Cardiometabolic risk factors lead to coronary endothelial and microvascular dysfunction that may cause chronotropic incompetence, or these factors may lead to premature cessation of exercise prior to reaching peak heart rate. This is an important distinction because if it is truly causal then improving chronotropy may result in lower cardiovascular risk; if it is simply a marker of premature cessation of exercise (i.e. coronary endothelial or mitochondrial dysfunction), then chronotropic incompetence may simply be a surrogate marker for subclinical cardiovascular disease. In the general population, there are strong associations between chronotropic incompetence at baseline and subsequent incident cardiovascular disease [10] including myocardial infarction [11], ischemic stroke [12], and mortality [10, 11, 13, 14]. This has not been studied prospectively among PWH, and whether improvement in chronotropy is associated with lower risk of cardiovascular events is unknown.

Several limitations of this secondary analysis should be noted. This was of an already completed clinical trial not designed to test the baseline prevalence of chronotropic incompetence among PWH or the effect of exercise training on chronotropy, which has distinct advantages and disadvantages. A small number of individuals had chronotropic incompetence. Individuals with known coronary artery disease and those taking chronotropic medications were included in the trial. For this secondary analysis, we chose not to exclude those on chronotropic medications but rather to use the lower threshold that has been previously published. A higher than expected proportion of participants had a low respiratory exchange ratio indicating a submaximal or VO_2_ peak instead of VO_2_max, likely due to deconditioning in this sedentary population. However, the foundational papers suggest that the adjusted heart rate reserve using the peak heart rate can still be used in submaximal tests because it is a ratio of the percentage of the heart rate reserve used to the metabolic reserve used and not affected by the exercise protocol, stage of exercise, or functional status [10, 13, 37].

In summary, we have found that chronotropic incompetence may contribute to reduced exercise capacity among PWH and improves with exercise training. Questions remain as to why PWH have a higher prevalence of chronotropic incompetence than the general population, and whether chronotropic incompetence is associated with subsequent cardiovascular events or subclinical cardiovascular disease among PWH. Taken in context of the numerous other benefits of exercise, we suggest that exercise can be recommended to individuals with chronotropic incompetence. Further studies are needed to explore the mechanisms for these improvements and the intensity of exercise training needed to modify the effect on chronotropy among PWH.

## Data Availability

Data is available upon reasonable request to the authors.

## References

1. Feinstein MJ, Hsue PY, Benjamin LA, et al. Characteristics, Prevention, and Management of Cardiovascular Disease in People Living With HIV: A Scientific Statement From the American Heart Association. Circulation 2019; 140:e98–e124.

2. Pothoff G, Wassermann K, Ostmann H. Impairment of exercise capacity in various groups of HIV-infected patients. Respiration 1994; 61:80–5.

3. Oursler KK, Sorkin JD, Smith BA, Katzel LI. Reduced aerobic capacity and physical functioning in older HIV-infected men. AIDS Res Hum Retroviruses 2006; 22:1113–21.

4. Duong M, Dumas JP, Buisson M, et al. Limitation of exercise capacity in nucleoside-treated HIV-infected patients with hyperlactataemia. HIV Med 2007; 8:105–11.

5. Gomes Neto M, Conceicao CS, Ogalha C, Brites C. Aerobic capacity and health-related quality of life in adults HIV-infected patients with and without lipodystrophy. Braz J Infect Dis 2016; 20:76–80.

6. de Lima LRA, Silva DAS, da Silva KS, Pelegrini A, de Carlos Back I, Petroski EL. Aerobic Fitness and Moderate to Vigorous Physical Activity in Children and Adolescents Living with HIV. Pediatr Exerc Sci 2017; 29:377–87.

7. Orton PM, Sokhela DG, Nokes KM, Perazzo JD, Webel AR. Factors related to functional exercise capacity amongst people with HIV in Durban, South Africa. Health SA 2021; 26:1532.

8. De Lorenzo A, Meirelles V, Vilela F, Souza FC. Use of the exercise treadmill test for the assessment of cardiac risk markers in adults infected with HIV. J Int Assoc Provid AIDS Care 2013; 12:110–6.

9. Durstenfeld MS, Peluso MJ, Spinelli MA, et al. Association of SARS-CoV-2 infection and Cardiopulmonary Long COVID with Exercise Capacity and Chronotropic Incompetence among People with HIV. J Am Heart Assoc 2023.

10. Lauer MS, Okin PM, Larson MG, Evans JC, Levy D. Impaired heart rate response to graded exercise. Prognostic implications of chronotropic incompetence in the Framingham Heart Study. Circulation 1996; 93:1520–6.

11. Elhendy A, Mahoney DW, Khandheria BK, Burger K, Pellikka PA. Prognostic significance of impairment of heart rate response to exercise: impact of left ventricular function and myocardial ischemia. J Am Coll Cardiol 2003; 42:823–30.

12. Jae SY, Heffernan K, Kurl S, et al. Chronotropic Response to Exercise Testing and the Risk of Stroke. Am J Cardiol 2021; 143:46–50.

13. Lauer MS, Francis GS, Okin PM, Pashkow FJ, Snader CE, Marwick TH. Impaired chronotropic response to exercise stress testing as a predictor of mortality. JAMA 1999; 281:524–9.

14. Gulati M, Shaw LJ, Thisted RA, Black HR, Bairey Merz CN, Arnsdorf MF. Heart rate response to exercise stress testing in asymptomatic women: the st. James women take heart project. Circulation 2010; 122:130–7.

15. Reddy YNV, Koepp KE, Carter R, et al. Rate-Adaptive Atrial Pacing for Heart Failure With Preserved Ejection Fraction: The RAPID-HF Randomized Clinical Trial. JAMA 2023; 329:801–9.

16. Keteyian SJ, Brawner CA, Schairer JR, et al. Effects of exercise training on chronotropic incompetence in patients with heart failure. Am Heart J 1999; 138:233–40.

17. Coman J, Freedman R, Koplan BA, et al. A blended sensor restores chronotropic response more favorably than an accelerometer alone in pacemaker patients: the LIFE study results. Pacing Clin Electrophysiol 2008; 31:1433–42.

18. Erlandson KM, MaWhinney S, Wilson M, et al. Physical function improvements with moderate or high-intensity exercise among older adults with or without HIV infection. AIDS 2018; 32:2317–26.

19. Khan MN, Pothier CE, Lauer MS. Chronotropic incompetence as a predictor of death among patients with normal electrograms taking beta blockers (metoprolol or atenolol). Am J Cardiol 2005; 96:1328–33.

20. Dobre D, Zannad F, Keteyian SJ, et al. Association between resting heart rate, chronotropic index, and long-term outcomes in patients with heart failure receiving β-blocker therapy: data from the HF-ACTION trial. Eur Heart J 2013; 34:2271–80.

21. Jankowski CM, Mawhinney S, Wilson MP, et al. Body Composition Changes in Response to Moderate-or High-Intensity Exercise Among Older Adults With or Without HIV Infection. Jaids-J Acq Imm Def 2020; 85:340–5.

22. Craig CL, Marshall AL, Sjöström M, et al. International physical activity questionnaire: 12-country reliability and validity. Med Sci Sports Exerc 2003; 35:1381–95.

23. Erlandson KM, Wilson MP, MaWhinney S, et al. The Impact of Moderate or High-Intensity Combined Exercise on Systemic Inflammation Among Older Persons With and Without HIV. J Infect Dis 2021; 223:1161–70.

24. Durstenfeld MS PM, Spinelli MA, Li D, Sander E, Swaminathan S, Arechiga VM, Hoh R, Aras MA, Long CS, Deeks SG, Hsue PY. EXERCISE CAPACITY IS REDUCED IN HIV INDEPENDENT OF SARS-COV-2 INFECTION In: CROI 2023. (Seattle, WA).

25. Kawasaki T, Kaimoto S, Sakatani T, et al. Chronotropic incompetence and autonomic dysfunction in patients without structural heart disease. Europace 2010; 12:561–6.

26. Brubaker PH, Kitzman DW. Chronotropic incompetence: causes, consequences, and management. Circulation 2011; 123:1010–20.

27. Hsue PY, Hunt PW, Ho JE, et al. Impact of HIV infection on diastolic function and left ventricular mass. Circ Heart Fail 2010; 3:132–9.

28. Sinha A, Ma Y, Scherzer R, et al. Role of T-Cell Dysfunction, Inflammation, and Coagulation in Microvascular Disease in HIV. J Am Heart Assoc 2016; 5.

29. Butler J, Greene SJ, Shah SH, et al. Diastolic Dysfunction in Patients With Human Immunodeficiency Virus Receiving Antiretroviral Therapy: Results From the CHART Study. J Card Fail 2020; 26:371–80.

30. Durstenfeld MS, Peluso MJ, Kaveti P, et al. Reduced Exercise Capacity, Chronotropic Incompetence, and Early Systemic Inflammation in Cardiopulmonary Phenotype Long Coronavirus Disease 2019. J Infect Dis 2023; 228:542–54.

31. Tseng ZH, Moffatt E, Kim A, et al. Sudden Cardiac Death and Myocardial Fibrosis, Determined by Autopsy, in Persons with HIV. N Engl J Med 2021; 384:2306–16.

32. Hansen D, Dendale P. Modifiable predictors of chronotropic incompetence in male patients with type 2 diabetes. J Cardiopulm Rehabil Prev 2014; 34:202–7.

33. Franssen WMA, Keytsman C, Marinus N, et al. Chronotropic incompetence is more frequent in obese adolescents and relates to systemic inflammation and exercise intolerance. J Sport Health Sci 2023; 12:194–201.

34. Huang PH, Leu HB, Chen JW, et al. Comparison of endothelial vasodilator function, inflammatory markers, and N-terminal pro-brain natriuretic peptide in patients with or without chronotropic incompetence to exercise test. Heart 2006; 92:609–14.

35. Lauer MS, Pashkow FJ, Larson MG, Levy D. Association of cigarette smoking with chronotropic incompetence and prognosis in the Framingham Heart Study. Circulation 1997; 96:897–903.

36. Srivastava R, Blackstone EH, Lauer MS. Association of smoking with abnormal exercise heart rate responses and long-term prognosis in a healthy, population-based cohort. Am J Med 2000; 109:20–6.

37. Wilkoff BL, Miller RE. Exercise testing for chronotropic assessment. Cardiology clinics 1992; 10:705–17.

